# Left Carotid Bulb Intima-Media Thickness Correlates with Age and Specific T lymphocyte Populations in People Living with and without HIV

**DOI:** 10.1101/2025.08.01.25332791

**Authors:** Lweendo Muchaili, Joreen P. Povia, Benson M. Hamooya, John R Koethe, Annet Kirabo, Sepiso K. Masenga

## Abstract

**Background:** Cardiovascular disease (CVD) is a leading cause of mortality worldwide, with carotid atherosclerosis playing a key role in its progression. Carotid intima-media thickness (CIMT) is a well-established biomarker for subclinical atherosclerosis. Emerging evidence suggests that immune dysregulation, particularly T-cell activation and exhaustion, may contribute to vascular pathology. People living with HIV (PLWH) face an elevated risk of CVD, yet the relationship between immune markers and CIMT in this population remains unclear.

**Methodology:** This cross-sectional study analyzed 100 participants, 70 virologically suppressed PLWH on antiretroviral therapy and 30 HIV-negative controls. Left carotid bulb intima-media thickness (LCBIMT) was measured via ultrasound, and immune markers (CD4+/CD8+ subsets, PD-1+ T-cells, IL-2, IL-17) were assessed. Statistical analyses included Spearman correlations and linear regression models stratified by HIV status to evaluate associations between LCBIMT, age, and lymphocyte subsets.

**Results:** Age was significantly associated with LCBIMT in both PLWH (β=0.0051, p<0.01) and controls (β=0.0101, p=0.001) groups. CD4+ intermediate % independently predicted LCBIMT in both populations (PLWH: β=0.0110, p=0.02; controls: β=0.0400, p<0.01). Notably, CD4+ naïve % showed an unexpected positive association in control participants (β=0.0050, p=0.03). T-cell exhaustion markers (PD-1+) were attenuated after adjustment, while CD8+ intermediate % approached significance in the controls (β=-0.0112, p=0.05).

**Conclusion:** HIV infection modifies cardiovascular risk through distinct immune mechanisms, particularly T-cell exhaustion and subset redistribution. These findings suggest immune profiling may aid in early CVD risk stratification and identify potential targets for intervention in high-risk populations, including PLWH.

## Introduction

Cardiovascular disease (CVD) remains a leading cause of morbidity and mortality worldwide, responsible for approximately 17.9 million deaths per year [1]. Most CVDs are associated with hypertension, carotid arterial wall remodeling, and atherosclerosis [2]. According to the World Health Organization (WHO), hypertension affects an estimated 1.28 billion people globally [3]. More than 60% of the individuals with hypertension have carotid plaques, a key feature of atherosclerosis [4,5]. Individuals with carotid atherosclerosis are at high risk of carotid artery stenosis, with 24% of asymptomatic stenotic patients dying within the first 5 years [6].

People living with HIV (PLWH) face a heightened vulnerability to cardiovascular disease (CVD) despite effective antiretroviral therapy (ART), exhibiting accelerated atherosclerosis and increased risk of cardiovascular events compared to the general population [7]. This elevated risk persists even with viral suppression, suggesting that chronic immune activation, persistent inflammation, and ART-related metabolic disturbances contribute to endothelial dysfunction and vascular remodeling [8]. Notably, carotid artery pathology in PLWH demonstrates distinct features compared to HIV-negative individuals, including more diffuse atherosclerotic lesions showing predilection for the carotid bulb, increased arterial stiffness, and unique patterns of immune cell infiltration [9]. While traditional risk factors like age and hypertension influence carotid intima-media thickness (CIMT) in both populations, PLWH exhibit stronger associations with T-cell exhaustion markers and intermediate monocyte activation, reflecting HIV-specific immunopathogenic mechanisms [10–12]. These differences highlight the need for tailored cardiovascular risk assessment strategies in HIV care, as conventional prediction models may underestimate risk in this population. Understanding the interplay between chronic HIV infection, immune dysregulation, and carotid atherosclerosis could provide critical insights for early intervention and targeted therapies to mitigate CVD risk in PLWH.

Early diagnosis of carotid pathology is important in the prevention of CVD associated with carotid atherosclerosis[13]. The Carotid intima-media thickness (CIMT) serves as a reliable, non-invasive diagnostic feature for assessing subclinical atherosclerosis and provides valuable prognostic information on CVD risk [14]. Measurements at the carotid bulb, where the artery bifurcates, offer valuable insights due to this region’s heightened susceptibility to hemodynamic stress and inflammation-driven plaque formation. This accelerated CIMT progression is of utmost relevance in PLWH, where chronic inflammation exacerbates atherogenesis. An increase in CIMT is an indicator of pathology [15,16].

Emerging evidence suggests that immune cells are significantly involved in cardiovascular pathology, especially inflammation-driven atherosclerosis. CD4+ T helper cells have been implicated in promoting vascular inflammation, which in turn speeds up the progression of atherosclerotic lesions. However, the exact nature of this relationship and how lymphocyte subsets correlate with CIMT remain poorly understood [17].

This study investigated the relationship between left carotid bulb intima-media thickness (LCBIMT), age, lymphocyte subsets, and immune markers in both PLWH and HIV-negative individuals. By examining these associations, we sought to clarify whether specific immune profiles, particularly those related to T-cell activation and exhaustion, may serve as markers of subclinical atherosclerosis in the context of HIV infection. This research has potential implications for early detection of vascular changes in high-risk populations, including PLWH, and could provide a basis for targeted interventions to modulate immune-related risk factors in CVD.

## Methodology

We utilized data from the Adiposity and Immune Activation Cohort of 100 individuals recruited at the Vanderbilt Comprehensive Care Clinic and the Vanderbilt University Medical Center Internal Medicine clinics between 2013 and 2014. The 70 PLWH participants recruited from the VCCC were evenly distributed across four BMI categories: <25.0, 25.0–29.9, 30.0–34.9, and ≥35.0 kg/m^2^. Each BMI stratum included similar proportions of males and females, as well as white and non-white individuals. All participants with HIV had been on a stable regimen of efavirenz, tenofovir, and emtricitabine (the combination pill *Atripla*) for at least six months before enrollment and had maintained undetectable HIV-1 RNA levels (<50 copies/mL) for the preceding two years. Additional inclusion criteria included a CD4+ count >350 cells/µL at enrollment, no use of anti-diabetic or statin medications in the prior six months, no heavy alcohol consumption (>11 drinks/week) or cocaine/amphetamine use, no active infections other than HIV, and no history of diabetes, cardiovascular disease (CVD), or rheumatologic conditions.

Thirty, HIV-negative controls were recruited from the Vanderbilt University Medical Center Internal Medicine clinics, matched by sex and race to the HIV-infected group, and distributed equally across BMI categories of 30.0–34.9 and ≥35.0 kg/m^2^. Controls had no history of anti-diabetic or statin use, no alcohol or illicit drug abuse, and no active infections or chronic conditions such as diabetes, CVD, or rheumatologic diseases.

Data on ART history, CD4+ counts, and viral loads were obtained from medical records, while smoking status was self-reported. All 100 participants underwent the same clinical research evaluation at the Vanderbilt Clinical Research Center after a minimum 8 hour fast. Brachial artery reactive hyperemia and CIMT were measured using a Philips iE33 ultrasound. Flow-mediated dilation (FMD) was calculated as the maximum percent increase in brachial artery diameter after cuff deflation. CIMT was measured at plaque-free segments of the carotid bulb and proximal internal carotid artery (ICA) using EKG-gated imaging.

Fasting blood samples were collected to measure high-sensitivity C-reactive protein (hsCRP), glucose, and biomarkers of immune activation (soluble CD14, CD163, IL-6, TNF-α receptor 1, ICAM-1, and VCAM-1) using ELISA and immunoassays. Anthropometric measurements were performed in triplicate.

### Immunophenotyping and Cell Subset Definitions

#### Naïve T cells

Naïve CD4+ and CD8+ T cells are antigen-inexperienced T lymphocytes that have not yet encountered their cognate antigen. They are characterized by the co-expression of CD45RA and CCR7, along with high expression of CD62L and CD27, and low or absent expression of activation markers CD38, HLA-DR, and checkpoint inhibitors such as PD-1.

#### Intermediate T cells

Intermediate CD4+ and CD8+ T cells are transitional phenotypes situated between naïve and fully differentiated memory or effector T cells.

Intermediate CD4+ T cells display a phenotype that includes partial down-regulation of naïve markers such as CD45RA and CCR7, with concurrent up-regulation of activation or memory-associated markers such as CD38, HLA-DR, CD69, PD-1, CD27, and CD28. They may also exhibit transitional co-stimulatory or inhibitory profiles, including intermediate expression of ICOS, CD25, or CTLA-4.

Intermediate CD8+ T cells are characterized by partial effector differentiation, marked by moderate or heterogeneous expression of CD45RA, CCR7, CD27, and CD28, alongside activation markers such as CD38, HLA-DR, and inhibitory receptors including PD-1, TIM-3, LAG-3, and TIGIT.

#### Statistical analyses

Statistical analyses were performed using StatCrunch software. The Shapiro-Wilk test was employed to assess the normality of the data distribution. To evaluate the associations between demographic, immunological, and inflammatory markers with left carotid bulb intima-media thickness (LCBIMT), Spearman correlation analysis was conducted, as the data did not meet the assumptions of normality. Additionally, simple and multiple linear regression analyses were performed to further explore the relationships between the variables and LCBIMT. All statistical tests were two-tailed, and a *p*-value of <0.05 was considered statistically significant.

#### Study approval

The study was approved by the Vanderbilt University Medical Center Institutional Review Board. All participants gave written informed consent. The research team conducted the study per the guidelines set by the United States Department of Health and Human Services. The trial is registered on ClinicalTrials.gov under the identifier NCT04439448.

## Results

### Characteristics of Study Participants

The study population consisted of 70 PLWH and 30 HIV-negative controls. Among the HIV-negative, there was a female predominance (60%, n=18) compared to males (40%, n=12), while PLWH showed a higher proportion of males (57%, n=40) than females (43%, n=30). Racial distribution differed between groups, with controls consisting of African American (37%, n=11) and Caucasian (63%, n=19) participants. PLWH included African American (49%, n=34), Caucasian (46%, n=32), Asian (1%, n=1), Hispanic (3%, n=2), and other racial groups (1%, n=1).

PLWH had higher median IL-2 levels (0.6 pg/mL) compared to controls (0.1985 pg/mL), and slightly elevated IL-17 levels (58.3 pg/mL vs 55.95 pg/mL). PD-1 expression was more pronounced in PLWH for both CD4+ (39% vs 30%) and CD8+ T cells (35.6% vs 25%). Naïve T cell proportions were lower in PLWH for both CD4+ (85% vs 90%) and CD8+ subsets (41% vs 54%).

Clinical measurements showed PLWH had marginally higher left carotid bulb intima-media thickness (0.65 mm vs 0.61 mm) and were older (median 45 years vs 37 years) but had lower BMI (30.85 vs 36.3) compared to controls (Table 1).

**Table 1:** Demographic and Clinical characteristics of study participants **Characteristic** HIV-negative (n = 30) PLWH (n = 70) **Sex, n (%)**

| Characteristic | HIV-negative (n = 30) | PLWH (n = 70) |
| --- | --- | --- |
| Sex, n (%) |  |  |
| Male | 12 (40.0) | 40 (57) |
| Female | 18 (60.0) | 30 (43) |
| **Race/Ethnicity, n (%)** |  |  |
| African American | 11 (37) | 34 (49) |
| Caucasian | 19 (63) | 32 (46) |
| Asian | 0 (0.0) | 1 (1) |
| Hispanic | 0 (0.0) | 2 (3) |
| Other | 0 (0.0) | 1 (1) |
| **Age (years), median (IQR)** | 37 (16) | 45 (11) |
| **BMI (kg/m<sup>2</sup>), median (IQR)** | 36.3 (8.6) | 30.85 (11.4) |
| **Inflammatory Markers, median (IQR)** |  |  |
| IL-2 (pg/mL) | 0.1985 (0.635) | 0.6 (0.514) |
| IL-17 (pg/mL) | 55.95 (42.8) | 58.3 (51.7) |
| **T Cell Activation and Differentiation, median (IQR)** |  |  |
| CD4<sup>+</sup> PD-1<sup>+</sup> (%) | 30.05 (14.5) | 39.1 (16.9) |
| CD8<sup>+</sup> PD-1<sup>+</sup> (%) | 25.25 (12.6) | 35.6 (16.2) |
| CD4<sup>+</sup> Naïve (%) | 90.1 (9.12) | 85.1 (21.56) |
| CD4<sup>+</sup> Intermediate (%) | 1.83 (2.28) | 1.67 (1.89) |
| CD8<sup>+</sup> Naïve (%) | 54.48 (46.99) | 40.73 (38.95) |
| CD8<sup>+</sup> Intermediate (%) | 7.57 (6.98) | 7.43 (5.56) |
| **Left carotid bulb IMT (mm), median (IQR)** | 0.61 (0.18) | 0.65 (0.155) |

*Abbreviations: PLWH – People living with HIV; IMT – Intima-media thickness; IL – Interleukin; PD-1 –* *Programmed cell death protein 1; BMI – Body mass index.*

### Spearman’s correlation analysis between Left carotid bulb intima thickness and independent variables

#### Control Population

Age and CD4 intermediate percentage had a significant association with LCBMIT. Age showed a moderate positive correlation with LCBMIT (β=0.4152, p=0.02). CD4+ intermediate % demonstrated the strongest association (β=0.4589, p=0.012, while CD4+ naïve % showed a negative but non-significant trend (β=-0.3236, p=0.08). No significant correlations were found for sex, race, BMI, IL-2, IL-17, or PD-1+ T-cell subsets (Figure 1).

**Figure 1:**
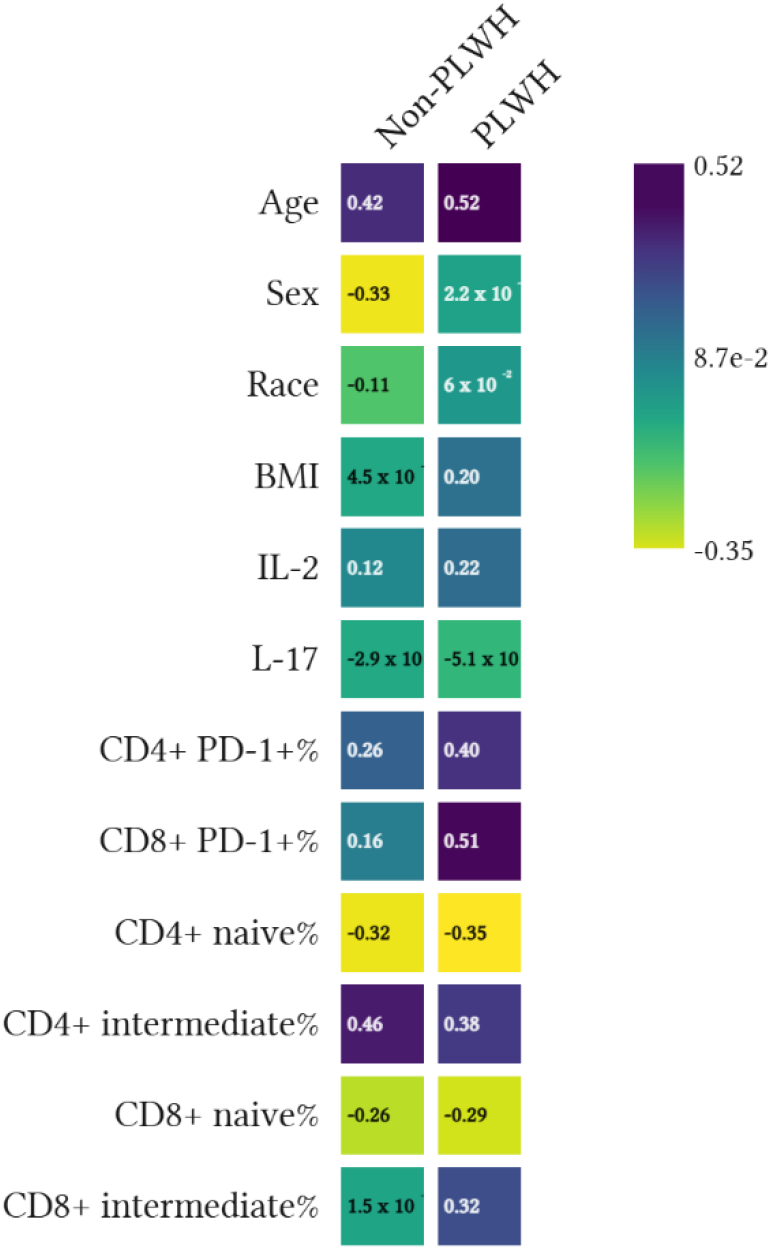
Spearman’s correlation analysis between LCBMIT and independent variables.

#### HIV Population

Age strongly correlated with LCBMIT (β=0.5224, p<0.001). Markers of T-cell exhaustion showed robust associations (CD8+ PD-1+%: β=0.5079, p<0.001; CD4+ PD-1+%: β=0.3973, p=0.001). Intermediate T-cell subsets were positively correlated while naïve subsets showed negative correlations, mirroring patterns seen in the control group.

### Simple linear regression analysis of left carotid bulb intima thickness and independent variables

#### Control Population

Age remained significant (β=0.0087, p=0.0007). Females showed lower CBMIT values (β=-0.1197, p=0.04). CD4+ intermediate % was non-significant (β=0.0190, p=0.10) while CD8+ naïve % showed borderline protection (β=-0.0022, p=0.05). Refer to figure 1 and 2 below

**Figure 2:**
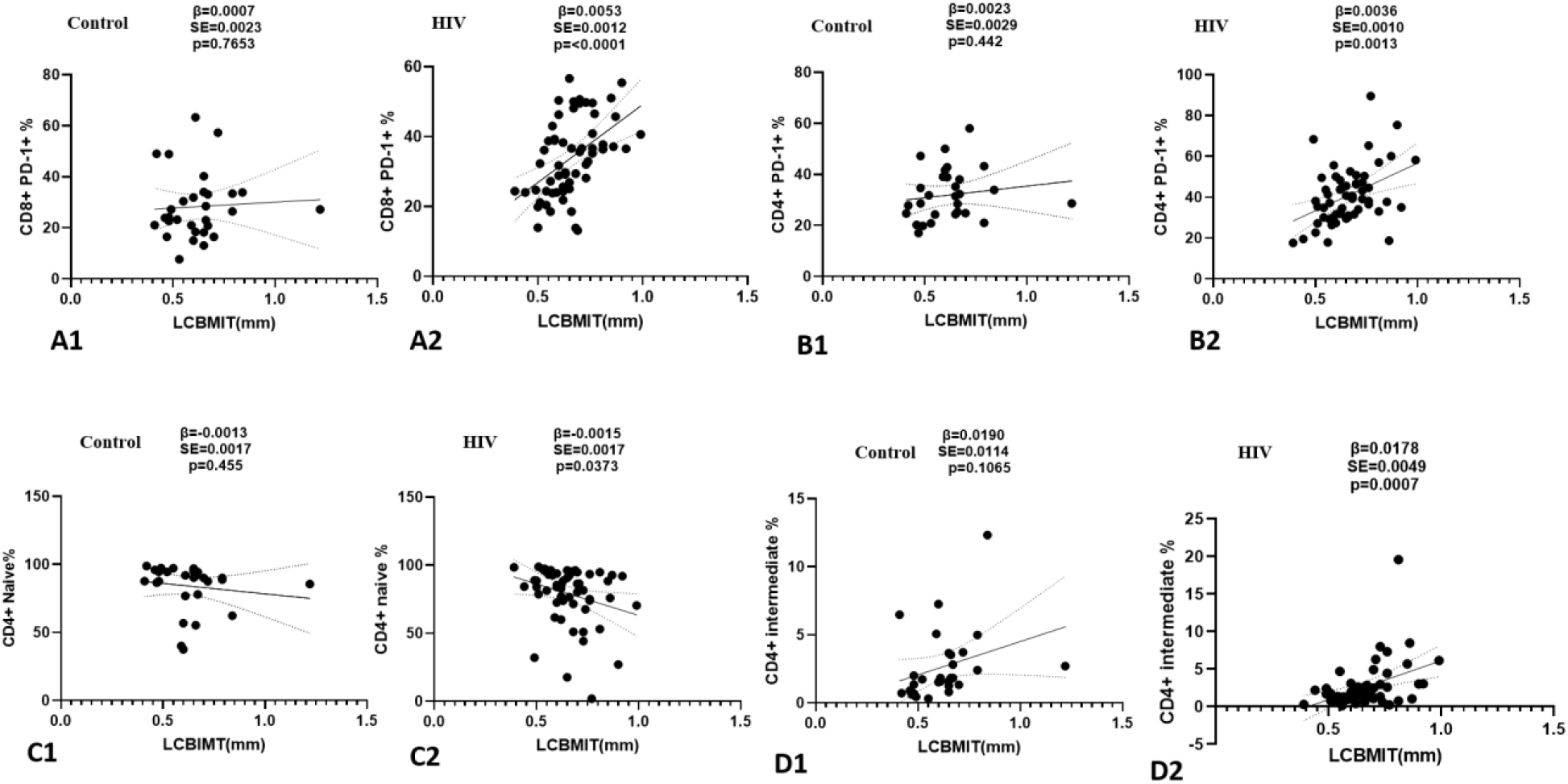
Simple linear regression between CBMIT and the independent variables for the control population. 2A1-D2 illustrates the magnitude and direction of association between demographic (Age) and immune variables (CD4+ PD-1+ %, CD8+ PD-1+ %, naïve and intermediate T-cell subsets) and CBMIT using standardized beta coefficients from multiple linear regression models among controls participants. Bars extending to the right indicate positive associations, while those extending to the left indicate negative associations. Error bars represent 95% confidence intervals. Abbreviations: **LCBMIT**-Left carotid bulb medial intima-media thickness; **PLWH:** people living with HIV.

**Figure 3:**
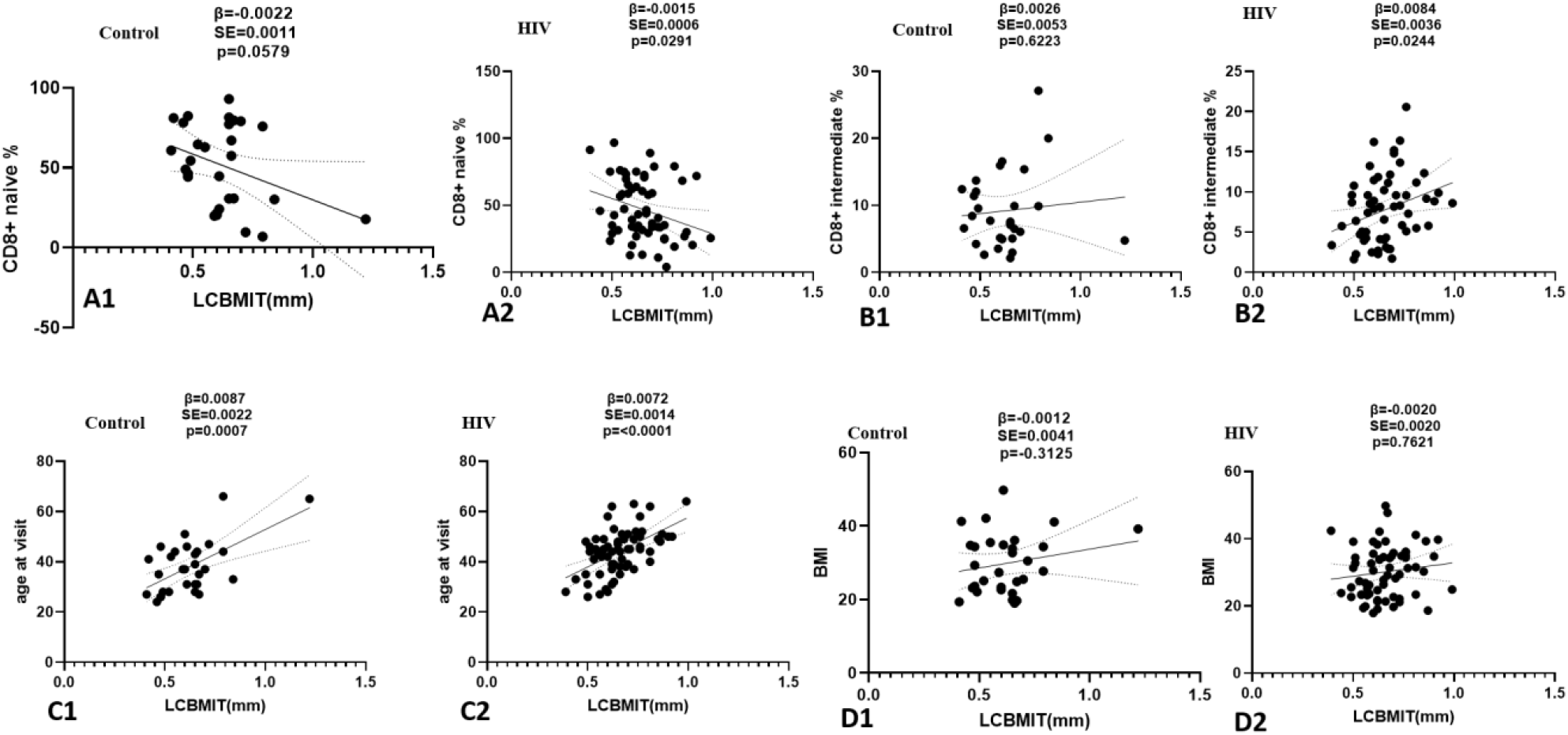
Simple linear regression between CBMIT and the independent variables for the control population. 3A1— D2 illustrates the magnitude and direction of association between demographic (Age) and immune variables (CD4+ PD-1+ %, CD8+ PD-1+ %, naïve and intermediate T-cell subsets) and CBMIT using standardized beta coefficients from multiple linear regression models among controls participants. Bars extending to the right indicate positive associations, while those extending to the left indicate negative associations. Error bars represent 95% confidence intervals. Abbreviations: **LCBMIT**-Left carotid bulb medial intima-media thickness; **PLWH:** people living with HIV.

**Figure 4:**
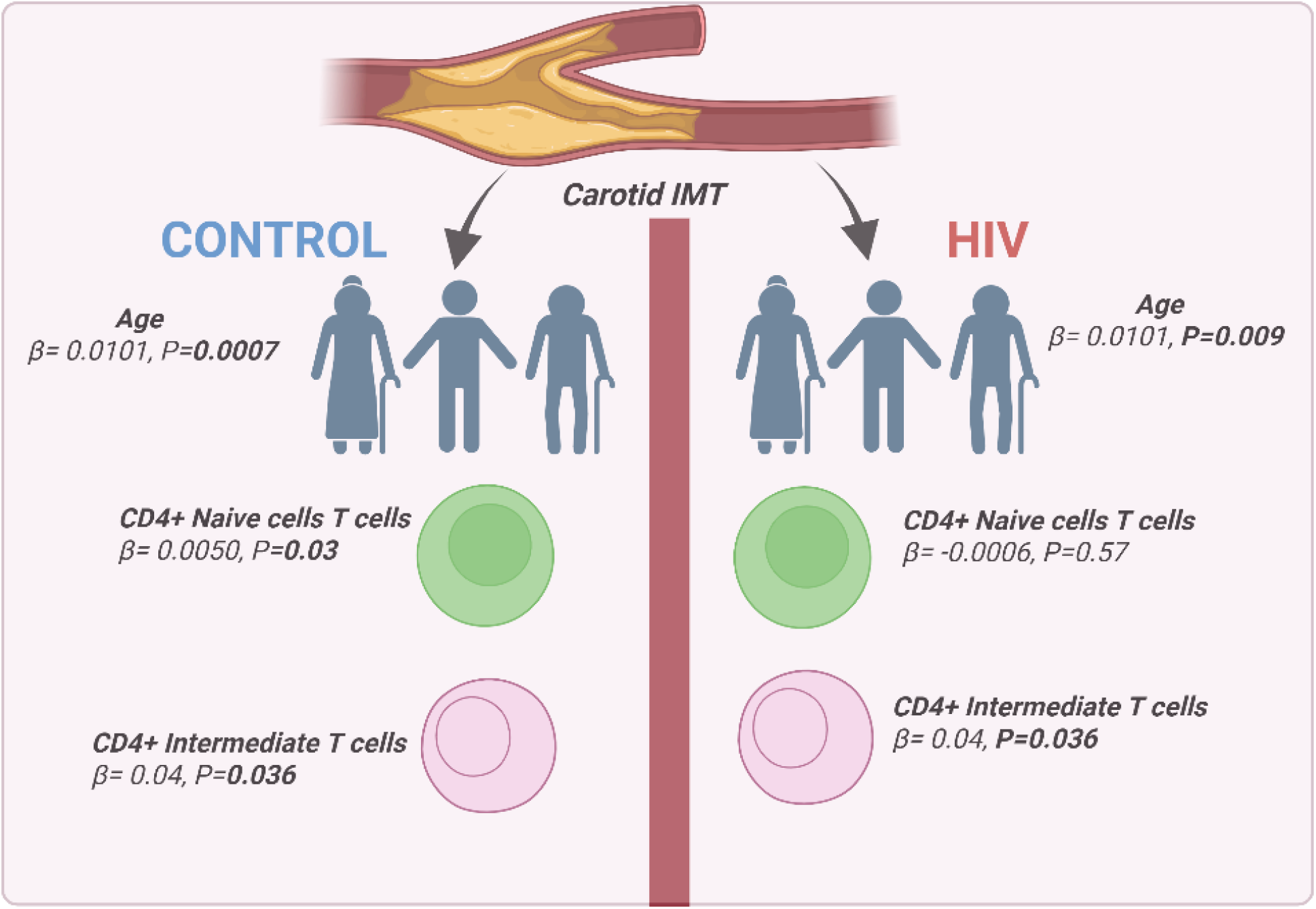
Multiple linear regression analysis highlights differing predictors of carotid intima-media thickness (CIMT) between HIV-positive individuals and HIV-negative controls. In controls, CIMT was significantly associated with age, CD4+ naïve T cells, and intermediate T cells. However, in people living with HIV (PLWH), while age and intermediate T cells remained significant predictors, CD4+ naïve T cells lost their association with CIMT

#### HIV Population

Age was a strong independent predictor (β=0.007, p<0.0001). PD-1+ T-cell subsets showed significant associations (CD8+: β=0.005, p<0.001; CD4+: β=0.003, p=0.001). CD4+ intermediate % had the largest effect size (β=0.0178, p=0.0007). Naïve T-cells demonstrated protective effects (CD4+: β=-0.0015, p=0.037; CD8+: β=-0.0015, p=0.029).

### Multiple linear regression analysis of left carotid bulb intima thickness and independent variables

#### Control Population

Age (β=0.0101, p=0.0007) and CD4+ intermediate % (β=0.0400, p=0.0036) remained strong predictors, Table 2. CD4+ naïve % unexpectedly showed positive association (β=0.0050, p=0.03). CD8+ intermediate % approached significance (β=-0.0112, p=0.0528).

**Table 2:** shows the multiple linear regression between CBMIT and the independent variables (control population)

| Variable | Beta | Standard Error | p-value | 95% CI |
| --- | --- | --- | --- | --- |
| Age at visit | 0.0101 | 0.0025 | **0.0007** | 0.0048 to 0.0155 |
| Sex | -0.0167 | 0.0511 | 0.7466 | -0.1234 to 0.0899 |
| CD4+ PD-1+ % | -0.0005 | 0.0035 | 0.8909 | -0.0079 to 0.0069 |
| CD8+ PD-1+ % | -0.0018 | 0.0025 | 0.4743 | -0.0071 to 0.0034 |
| CD4+ naïve % | 0.0050 | 0.0021 | **0.0300** | 0.0005 to 0.0095 |
| CD4+ intermediate % | 0.0400 | 0.0121 | **0.0036** | 0.0147 to 0.0655 |
| CD8+ naïve % | -0.0023 | 0.0014 | 0.1218 | -0.0053 to 0.0007 |
| CD8+ intermediate % | -0.0113 | 0.0054 | 0.0528 | -0.0227 to 0.0001 |

#### PLWH Population

Age (β=0.0051, p=0.009) and CD4+ intermediate % (β=0.0110, p=0.02) maintained significance, Table 3. PD-1+ associations were attenuated after adjustment.

**Table 3:** multiple linear regression between CBMIT and the independent variables (PLWH population)

|  |  |  |  |  |
| --- | --- | --- | --- | --- |
| Variable | Beta | Standard Error | p-value | 95% CI |
| Age at visit | 0.0051 | 0.0019 | **0.009** | 0.0013 to 0.0089 |
| Sex | -0.0167 | 0.0316 | 0.59 | -0.0801 to 0.0467 |
| CD4+ PD-1+ % | 0.0010 | 0.0016 | 0.50 | -0.0021 to 0.0042 |
| CD8+ PD-1+ % | 0.0025 | 0.0017 | 0.14 | -0.0009 to 0.0059 |
| CD4+ naïve % | -0.0006 | 0.0010 | 0.57 | -0.0026 to 0.0014 |
| CD4+ intermediate % | 0.0110 | 0.0047 | **0.02** | 0.0016 to 0.0204 |
| CD8+ naïve % | 0.0007 | 0.0008 | 0.36 | -0.0009 to 0.0023 |
| CD8+ intermediate % | 0.0009 | 0.0037 | 0.81 | -0.0067 to 0.0084 |

## Discussion

Our findings provide compelling evidence that immune dysregulation contributes to subclinical atherosclerosis through distinct pathways in PLWH versus HIV-negative controls. The multivariate regression analyses reveal several critical insights about cardiovascular risk stratification in these populations.

Age demonstrated consistent associations with CBMIT in both groups. In the control population, age at visit had the strongest effect size (β=0.0101, p=0.0007) among all significant predictors, indicating that each one-year increase in age was associated with a 0.0101-unit increase in CBMIT. In contrast, in the PLWH population, the effect size of age was approximately half as large (β=0.0051, p=0.0096). This suggests that while aging remains a significant factor in both groups, its impact on CBMIT is stronger in individuals with HIV. This is consistent with the literature, as previous studies have shown that CIMT progression in PLWH does not follow a strongly age-predictable pattern, unlike in the general population [18,19].

The differential immune cell associations with CMIT observed between controls and PLWH populations reveal shared and distinct immunological pathways influencing subclinical atherosclerosis. Most interesting, CD4+ intermediate cells emerged as the strongest immune predictor of CIMT in controls (β=0.0400, p=0.003), with a persistent though attenuated effect in the HIV-positive group (β=0.0110, p=0.02). This consistency across populations suggests these transitional T-cells, positioned between naïve and memory states, may play a fundamental role in vascular homeostasis, potentially through their dual capacity for immune regulation and effector functions. Our findings extend prior work linking CD4+ subsets to cardiovascular risk by demonstrating their independence from traditional risk factors, including in HIV, where immune dysregulation is prominent [20–22].

The positive association of CD4+ naïve cells with CIMT in controls (β=0.0050, p=0.03) contrasts with some previous studies. There have been notable variations in study findings on the role of CD4+ naïve cells. While some studies suggest that CD4+ naïve cells have a protective role against atherosclerosis, other studies indicate that they also contribute to disease progression [17,23,24]. It is interesting that in the PLWH, however, this association was absent, possibly reflecting ART-mediated reconstitution or competing effects of chronic inflammation.

The lack of significant associations between PD-1+ T-cell subsets and CIMT in the multiple linear regression models for the PLWH population presents an interesting contrast to their presence in Spearman correlation and univariate analyses. While the adjusted models account for potential confounders, the disappearance of PD-1+ associations should not be interpreted as evidence against their biological relevance. Alp et al. in their research showed that statistical significance alone does not guarantee clinical relevance, as small effects may be meaningful biologically even if statistically marginal, especially in studies with limited sample sizes like ours [25]. The persistence of PD-1+ correlations in both simple regression and Spearman analyses suggests that these exhaustion markers may influence CIMT through pathways that become obscured when accounting for other variables, especially given our study’s small sample size. Therefore, these findings should be interpreted with these limitations in mind.

Our study also demonstrates distinct patterns of PD-1 associations with CIMT between PLWH and the HIV negative populations. In controls, the lack of significant associations between PD-1+ T-cells and CIMT across all analyses aligns with current understanding of PD-1 biology, where it primarily functions as an immune regulator without established direct involvement in cardiovascular pathogenesis. This contrasts with PLWH, where PD-1’s biological plausibility in cardiovascular risk through chronic immune activation is well-documented [26,27]. These differential findings highlight fundamentally distinct immune mechanisms contributing to vascular pathology in these populations, while the HIV negative population appear to follow conventional cardiovascular risk pathways, PLWH likely experience immune-mediated vascular effects where PD-1 plays a key role [28–30]. These findings emphasize the importance of population-specific considerations when evaluating immune markers of cardiovascular risk.

The clinical implications of these findings are substantial. The consistent identification of CD4+ intermediate subsets as independent associates of CIMT suggests they could serve as valuable biomarkers for atherosclerosis risk assessment. In HIV care, where conventional risk calculators often underestimate cardiovascular risk, incorporating immune parameters like intermediate cell percentages could significantly improve risk prediction. These findings support emerging paradigms that view atherosclerosis as, in part, an immunologic disorder and suggest potential therapeutic targets for intervention [31,32].

From a public health perspective, our results show the need for differentiated approaches to cardiovascular prevention. For PLWH, the findings reinforce that viral suppression alone may not fully mitigate cardiovascular risk, supporting calls for more aggressive monitoring and management of immune-mediated risk pathways [33]. The attenuated but persistent immune-CIMT associations in this group suggest that adjunctive anti-inflammatory strategies merit investigation as complements to traditional risk factor control. In the general population, the strong association of CD4+ intermediate cells with subclinical atherosclerosis opens new avenues for risk stratification and possibly targeted immunomodulation in high-risk individuals.

This study has several important strengths, including its rigorous phenotyping of both vascular and immune parameters, careful matching of PLWH and HIV-Negative controls, and comprehensive statistical adjustment for potential confounders. The use of multiple linear regression models allows us to identify independent predictors while controlling for interrelated variables, providing clearer insight into potential causal pathways.

However, certain limitations must be acknowledged. The cross-sectional design precludes the determination of temporal relationships between immune parameters and CIMT progression. The moderate sample size, while adequate for detecting moderate to large effects, may have limited power to identify smaller associations. Additionally, while we controlled for numerous potential confounders, residual confounding by unmeasured variables remains possible.

Future research directions should include longitudinal studies to establish whether changes in these immune parameters predict CIMT progression over time and investigation of whether interventions targeting intermediate monocytes can modify cardiovascular risk. Mechanistic studies exploring how these immune subsets contribute to endothelial dysfunction and vascular remodeling would further elucidate the pathophysiology suggested by our findings. Additionally, larger studies incorporating clinical endpoints could determine whether these immune-CIMT relationships translate to hard cardiovascular outcomes.

## Conclusion

Our findings identify CD4+ intermediate cells as a key immune correlate of subclinical atherosclerosis in both PLWH and controls, while revealing HIV-specific differences in PD-1+ T-cell associations and age-related vascular risk. These results underscore the need for immune-aware cardiovascular screening, particularly in PLWH who demonstrate accelerated vascular aging. The study provides a foundation for developing targeted immunomodulatory strategies to mitigate atherosclerosis risk based on distinct HIV-status pathways.

## Data Availability

All data will be available upon request from the corresponding author

## Conflict of Interest

The authors declare that the research was conducted in the absence of any commercial or financial relationships that could be construed as a potential conflict of interest.

## Author Contributions

SKM and LM conceived the Study. LM and SKM wrote the original draft. AK, JRK and JPP edited and revised the manuscript. All authors contributed to the article edits and approved the final manuscript.

## Funding

This work was supported by the Fogarty International Center of the National Institutes of Health, National Institute of Diabetes and Digestive and Kidney Diseases of the National Institutes of Health grants R01HL144941 (AK), 2D43TW009744 (SKM), R21TW012635 (AK and SKM) and the American Heart Association Award Number 24IVPHA1297559 https://doi.org/10.58275/AHA. 24IVPHA1297559.pc.gr.193866 (AK and SKM), National Institute of Allergy and Infectious Diseases grant K23 AI100700, National Center for Advancing Translational Sciences grant UL1 TR002243, the Tennessee Center for AIDS Research grant P30 AI110527, and National Heart, Lung, and Blood Institute grant K01HL130497. The funders had no role in study design, data collection and analysis, decision to publish, or preparation of the manuscript. Its contents are solely the responsibility of the authors and do not necessarily represent official views of the National Center for Advancing Translational Sciences or the National Institutes of Health.

## Notes

### Competing Interest Statement

The authors have declared no competing interest.

### Clinical Trial

The trial is registered on ClinicalTrials.gov under the identifier NCT04439448.

## References

1. Cardiovascular diseases [Internet]. [cited 2025 May 13]. Available from: https://www.who.int/health-topics/cardiovascular-diseases#tab=tab_1

2. Pewowaruk R, Korcarz C, Tedla Y, Mitchell C, Gepner AD. Carotid Artery Stiffness Mechanisms in Hypertension and their Association with Echolucency and Texture Features: The Multi-Ethnic Study of Atherosclerosis (MESA). Ultrasound Med Biol. 2022 Nov;48(11):2249–57.

3. WHO. Hypertension [Internet]. World Health Organization. 2023 [cited 2023 Apr 3]. Available from: https://www.who.int/news-room/fact-sheets/detail/hypertension

4. Long NP. STUDYING THE SITUATION OF CAROTID ATHEROSCLEROSIS OF HYPERTENSIVE PATIENTS IN THE NORTH OF BINH DINH PROVINE, VIETNAM. J Hypertens. 2021 Apr;39:e158.

5. Prisant LM, Zemel PC, Nichols FT, Zemel MB, Sowers JR, Carr AA, et al. Carotid plaque associations among hypertensive patients. Arch Intern Med. 1993 Feb 22;153(4):501–6.

6. Giannopoulos A, Kakkos S, Abbott A, Naylor AR, Richards T, Mikhailidis DP, et al. Long-term Mortality in Patients with Asymptomatic Carotid Stenosis: Implications for Statin Therapy. Eur J Vasc Endovasc Surg. 2015 Nov 1;50(5):573–82.

7. Paternò Raddusa MS, Marino A, Celesia BM, Spampinato S, Giarratana C, Venanzi Rullo E, et al. Atherosclerosis and Cardiovascular Complications in People Living with HIV: A Focused Review. Infect Dis Rep. 2024 Sep 1;16(5):846–63.

8. Kaplan RC, Hanna DB, Kizer JR. Recent Insights into Cardiovascular Disease (CVD) Risk Among HIV-Infected Adults. Curr HIV/AIDS Rep. 2016 Feb;13(1):44–52.

9. Hsue PY, Scherzer R, Hunt PW, Schnell A, Bolger AF, Kalapus SC, et al. Carotid Intima-Media Thickness Progression in HIV-Infected Adults Occurs Preferentially at the Carotid Bifurcation and Is Predicted by Inflammation. J Am Heart Assoc Cardiovasc Cerebrovasc Dis. 2012 Apr 24;1(2):jah3–e000422.

10. Obare LM, Temu T, Mallal SA, Wanjalla CN. Inflammation in HIV and Its Impact on Atherosclerotic Cardiovascular Disease. Circ Res. 2024 May 24;134(11):1515–45.

11. Blaauw MJT, Berrevoets MAH, Vos Wajw, Groenendijk AL, van Eekeren LE, Vadaq N, et al. Traditional Cardiovascular Risk Factors Are Stronger Related to Carotid Intima-Media Thickness Than to Presence of Carotid Plaques in People Living With HIV. J Am Heart Assoc. 2023 Oct 17;12(20):e030606.

12. Hmiel L, Zhang S, Obare LM, Santana MA de O, Wanjalla CN, Titanji BK, et al. Inflammatory and Immune Mechanisms for Atherosclerotic Cardiovascular Disease in HIV. Int J Mol Sci. 2024 Jan;25(13):7266.

13. Lee CJ, Park S. The Role of Carotid Ultrasound for Cardiovascular Risk Stratification beyond Traditional Risk Factors. Yonsei Med J. 2014 May 1;55(3):551–7.

14. Nezu T, Hosomi N, Aoki S, Matsumoto M. Carotid Intima-Media Thickness for Atherosclerosis. J Atheroscler Thromb. 2016;23(1):18–31.

15. Johri AM, Chitty DW, Matangi M, Malik P, Mousavi P, Day A, et al. Can carotid bulb plaque assessment rule out significant coronary artery disease? A comparison of plaque quantification by two- and three-dimensional ultrasound. J Am Soc Echocardiogr Off Publ Am Soc Echocardiogr. 2013 Jan;26(1):86–95.

16. Hulthe J, Wikstrand J, Emanuelsson H, Wiklund O, de Feyter PJ, Wendelhag I. Atherosclerotic changes in the carotid artery bulb as measured by B-mode ultrasound are associated with the extent of coronary atherosclerosis. Stroke. 1997 Jun;28(6):1189–94.

17. Saigusa R, Winkels H, Ley K. T cell subsets and functions in atherosclerosis. Nat Rev Cardiol. 2020 Jul;17(7):387–401.

18. Hanna DB, Guo M, Bůžková P, Miller TL, Post WS, Stein JH, et al. HIV Infection and Carotid Artery Intima-media Thickness: Pooled Analyses Across 5 Cohorts of the NHLBI HIV-CVD Collaborative. Clin Infect Dis. 2016 Jul 15;63(2):249–56.

19. Hsue PY, Lo JC, Franklin A, Bolger AF, Martin JN, Deeks SG, et al. Progression of Atherosclerosis as Assessed by Carotid Intima-Media Thickness in Patients With HIV Infection. Circulation. 2004 Apr 6;109(13):1603–8.

20. Martín P, Sánchez-Madrid F. T cells in cardiac health and disease. J Clin Invest. 135(2):e185218.

21. Ramos GC, van den Berg A, Nunes-Silva V, Weirather J, Peters L, Burkard M, et al. Myocardial aging as a T-cell–mediated phenomenon. Proc Natl Acad Sci. 2017 Mar 21;114(12):E2420–9.

22. Xia Y, Gao D, Wang X, Liu B, Shan X, Sun Y, et al. Role of Treg cell subsets in cardiovascular disease pathogenesis and potential therapeutic targets. Front Immunol [Internet]. 2024 Mar 15 [cited 2025 May 19];15. Available from: https://www.frontiersin.org/journals/immunology/articles/10.3389/fimmu.2024.1331609/full

23. Olson NC, Doyle MF, Jenny NS, Huber SA, Psaty BM, Kronmal RA, et al. Decreased Naive and Increased Memory CD4+ T Cells Are Associated with Subclinical Atherosclerosis: The Multi-Ethnic Study of Atherosclerosis. PLOS ONE. 2013 Aug 23;8(8):e71498.

24. Podolec J, Niewiara L, Skiba D, Siedlinski M, Baran J, Komar M, et al. Higher levels of circulating naïve CD8+CD45RA+ cells are associated with lower extent of coronary atherosclerosis and vascular dysfunction. Int J Cardiol. 2018 May 15;259:26–30.

25. Alp HH, Tran MTC, Markus C, Ho CS, Loh TP, Zakaria R, et al. Clinical vs. statistical significance: considerations for clinical laboratories. Clin Chem Lab Med. 2025 Apr 8;

26. Sinha A, Ma Y, Scherzer R, Hur S, Li D, Ganz P, et al. Role of T-Cell Dysfunction, Inflammation, and Coagulation in Microvascular Disease in HIV. J Am Heart Assoc. 2016 Dec 20;5(12):e004243.

27. Bowler S, Chew GM, Budoff M, Chow D, Mitchell BI, DʼAntoni ML, et al. PD-1+ and TIGIT+ CD4 T Cells Are Associated With Coronary Artery Calcium Progression in HIV-Infected Treated Adults. J Acquir Immune Defic Syndr 1999. 2019 May 1;81(1):e21–3.

28. Feinstein MJ, Hsue PY, Benjamin L, Bloomfield GS, Currier JS, Freiberg MS, et al. Characteristics, Prevention, and Management of Cardiovascular Disease in People Living With HIV: A Scientific Statement From the American Heart Association. Circulation. 2019 Jul 9;140(2):e98– 124.

29. Adhikary D, Barman S, Ranjan R, Stone H. A Systematic Review of Major Cardiovascular Risk Factors: A Growing Global Health Concern. Cureus. 14(10):e30119.

30. Triant VA, Lyass A, Hurley LB, Borowsky LH, Ehrbar RQ, He W, et al. Cardiovascular Risk Estimation Is Suboptimal in People With HIV. J Am Heart Assoc. 2024 May 21;13(10):e029228.

31. Abplanalp WT, Merten M, Dimmeler S. Straight to the Heart: T Cells That Specifically Target Cardiac Tissue. Circulation. 2022 Dec 20;146(25):1946–9.

32. Wang X, Zhou H, Liu Q, Cheng P, Zhao T, Yang T, et al. Targeting regulatory T cells for cardiovascular diseases. Front Immunol [Internet]. 2023 Feb 23 [cited 2025 May 19];14. Available from: https://www.frontiersin.org/journals/immunology/articles/10.3389/fimmu.2023.1126761/full

33. Triant VA. Cardiovascular Disease and HIV Infection. Curr HIV/AIDS Rep. 2013 Sep;10(3):199–206.

